# Clinical Uncertainty In Large Vessel Occlusion Ischemic Stroke (CULVO): An Intrarater And Interrater Agreement Study

**DOI:** 10.1101/2023.12.18.23300180

**Authors:** Jose Danilo B. Diestro, Robert Fahed, Abdelsimar T. Omar, Christine Hawkes, Eef J. Hendriks, Clare Angeli Enriquez, Muneer Eesa, Grant Stotts, Hubert Lee, Shashank Nagendra, Alexandre Poppe, Célina Ducroux, Timothy Lim, Karl Narvacan, Michael Rizutto, Afra Alfalahi, Hidehisa Nishi, Pragyan Sarma, Vladislav Ze’ev Itsekzon-Hayosh, Katrina Ignacio, William Boisseau, Eduardo Pimenta Ribeiro Pontes Almeida, Anass Benomar, Mohammed A. Almekhlafi, Genvieve Milot, Aviraj Satish Deshmukh, Kislay Kishore, Donatella Tampieri, Jeffrey Z. Wang, Abhilekh Srivastava, Daniel Roy, Federico Carpani, Nima Kashani, Claudia Candale-Radu, Nishita Singh, Maria Bres Bullrich, Robert Sarmiento, Ryan Muir, Carmen Parra-Farinas, Stephanie D. Reiter, Yan Deschaintre, Ravinder-Jeet Singh, Vivel Bodani, Aristeidis Katsanos, Ronit Agid, Atif Zafar, Vitor Mendes Pereira, Julian Spears, Thomas R. Marotta, Pascal Djiadeu, Sunjay Sharma, Forough Farrokhyar

## Abstract

**Background:** Limited research exists regarding the impact of neuroimaging modality on endovascular thrombectomy (EVT) decisions for late window large vessel occlusion (LVO) stroke cases.

**Purpose:** This study assesses whether perfusion CT imaging: 1) alters the proportion of recommendations for EVT, and 2) enhances the reliability of EVT decision-making compared to non-contrast CT and CT angiography.

**Materials and Methods:** We conducted an online survey using 30 patients drawn from an institutional database of 3144 acute stroke cranial CT scans. These cases were presented to 29 stroke or neurointerventional physicians from Canada across two sessions. Physicians evaluated each patient both with and without perfusion imaging and gave EVT recommendations. We used non-overlapping 95% confidence intervals and difference in agreement classification as criteria to suggest a difference between the Gwet AC1 statistics (κ_G_). Our outcomes were: 1) the proportion of EVT recommendations, and 2) interrater and intrarater agreement, with or without perfusion imaging.

**Results:** In the first round, 29 raters completed the assessment, with 28 finishing the second round. The percentage of EVT recommendations differed by 1.1% with or without perfusion imaging. However, individual decisions changed in 21.4% of cases, with 11.3% against EVT and 10.1% in favor. Interrater agreement (κG) among the 29 raters was similar between non-perfusion CT neuroimaging and perfusion CT neuroimaging (κG = 0.487; 95% CI 0.327, 0.647 and κG = 0.552; 95% CI 0.430, 0.675). The 95% CIs overlapped with moderate agreement in both. Intrarater agreement exhibited overlapping 95% CIs for all 28 raters. κG was either substantial or excellent (0.81-1) for 71.4% (20/28) of raters in both groups.

**Conclusion:** The difference in EVT recommendations is minimal with either neuroimaing protocol. Regarding agreement we found that use of automated CT perfusion images does not significantly impact the reliability of EVT decisions for late window LVO patients.

## Introduction

The efficacy of endovascular thrombectomy (EVT) for large vessel occlusion stroke patients has been demonstrated by randomized controlled trials, indicating that the mechanical removal of the clot leads to better clinical outcomes compared to best medical management. ^1–5^ Non-perfusion CT neuroimaging which consists of a non-contrast cranial computed tomography (NCCT) scan and a computed tomography angiogram (CTA) along with clinical findings are deemed sufficient to recommend EVT for large vessel occlusion stroke presenting within 6 hours of presumed stroke onset.

For the late window (6-24 hours) however, both the American Heart Association and Canadian Stroke Best Practices recommend that perfusion CT neuroimaging (NCCT, CTA and perfusion imaging) be utilized to allow for more nuanced patient selection. ^6,7^ These recommendations are driven by use of perfusion imaging in late-window EVT trials. Both DAWN and DEFUSE 3, used perfusion imaging to derive quantitative values for core infarct and excluded those that were considered to have a large core. By including a strict core size in their inclusion criteria, these studies hoped to minimize futile recanalization. New randomized trials that also include late-window large vessel occlusion stroke patients have demonstrated that even with a large core, those who underwent EVT still derived a significant benefit. ^8–10^ Hence, it appears that the objection to employing EVT for large core infarcts on the grounds of futility is unfounded. Further evidence for non-perfusion CT has recently come in from the MR CLEAN

LATE trial that did not employ perfusion imaging prior to randomizing patients in the late window.^11^ Lastly, CLEAR and SOLSTICE consortiums found no difference in clinical outcomes between with or without perfusion neuroimaging. ^12,13^

We performed an experimental survey among stroke neurologists and neurointerventionalists, encompassing a wide range of large vessel occlusion stroke cases. The study aimed to examine: 1) the difference in proportion of EVT recommendations with and without perfusion imaging and, 2) whether the use of perfusion CT neuroimaging improves the reliability among physicians in repeated surveys (intra-rater) and among different physicians and experience (interrater), compared to non-perfusion CT neuroimaging. Reliability and agreement are used interchangeably across the text. We use these terms to refer to the propensity of the raters to make the same decision about a case after removal of chance agreement.^14^

We hypothesize that the reliability of EVT decisions will be similar based on either neuroimaging modality.

## Methodology

The study was prepared in accordance with the Guidelines for Reporting Reliability and Agreement Studies (GRRAS).^15^

### Case Selection

All included studies were taken from an institutional database of patients undergoing in-house neuroimaging for symptoms of acute ischemic stroke. A team of clinical and research physicians 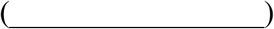 reviewed our database to identify reported large vessel occlusions in patients being scanned for symptoms of stroke from January 2018 to August 2022. __ a dual trained stroke neurologist and neurointerventionalist reviewed all the flagged studies to determine eligibility using the following criteria: 1) > 18 years old, 2) stroke onset or last known well time between 6-24 hours prior to start of imaging (late-window stroke), 3) use of automated perfusion CT (RAPID, iSchemaView, Menlo Park, CA), 4) confirmed large vessel occlusion on CTA involving: first segment of the middle cerebral artery (M1), terminal portion of the internal carotid artery or tandem occlusion, 5) penumbra size of at least 15cc of perfusion CT, 6) mismatch ratio of at least 1.8, 7) National Institutes of Health Stroke Score (NIHSS) of 6 or more. For quality control, neuroimaging with unsatisfactory elements (artifacts, improper timing of contrast, or poor perfusion study quality) were excluded.

To minimize the paradoxes of Kappa statistics,^16,17^ we included the full range of infarct core sizes: 10 cases with small core (0-49 cc), 10 cases with medium core (50-100cc), 10 cases with large core (>100cc). We selected cases with the intention of assessing the reliability (reproducibility) of clinician judgments within an experimental environment that encompasses a comprehensive range of core infarct sizes and clinical characteristics.^18^

### Rater Selection

We invited stroke neurologists and neurointerventionalists in Canada who work in EVT capable stroke centers with access to perfusion imaging. Clinical fellows with at least a year of fellowship were allowed to participate. Invitations were sent to the physicians through the Canadian Stroke Consortium and the Canadian Interventional Neuro Group mailing lists. The raters were classified as: stroke neurologists, neurointerventionalists (neuroradiologists and neurosurgeons) and dual-trained neurologists who are both stroke neurologists and neurointerventionalists.

### Survey

Selected cases were collected using Microsoft Excel v16.72 and placed into the survey format on REDCap (Research Electronic Data Capture). ^19,20^ Personalized links were emailed to the participants. The following case details were provided: age, location of occlusion, the NIHSS, time of onset, time of scan, non-perfusion CT neuroimaging (NCCT and CTA). The NCCT scans were shown in stroke window settings (window width of 35HU and window level of 35HU).^21–23^ The same 30 cases were shown again with the same information but with perfusion CT neuroimaging. Overall, the participants encountered 60 cases comprised of the same 30 patients shown twice with and without perfusion imaging. The raters were not informed that these 60 cases were the same 30 patients shown twice. To test intra-rater agreement, a second round of survey was done at least 3 weeks from the first. The survey was pilot tested and revised based on feedback from 7 trainee physicians who did not meet the study’s official rater criteria.

For each case, the raters were asked to grade ASPECTS, the single-phase collateral score (collateral score 0: absence of vessels distal to the occlusion, 1: ≤ 50% but >0% collateral supply, 2: ≥ 50% but <100% collateral supply or 3: 100% collateral supply) and finally whether they would recommend EVT based on the available clinical and radiologic data.^24,25^

### Sample Size

We estimated, using kappaSize package in R version 4.2.0 (R Foundation for Statistical Computing, Vienna, Austria), assuming an anticipated kappa value of 0.6 (substantial), that at least 30 cases were necessary for the lower limit of a 95% one-sided confidence interval to remain above 0.45 between at least 6 raters, considering an anticipated prevalence of EVT recommendation of 0.5.^26^ A rule of thumb for agreement studies with binary outcomes is to have at least 10 raters reviewing at least 30-50 patients.^18^ While more patients will enable us to provide an even wider spectrum of patients, we also want to avoid rater fatigue. For this study, we selected 60 cases with three questions each. We needed at least 6 raters based on the sample size computation but aimed for at least 10 to follow the current convention.

### Agreement Statistics

The inter-rater and intra-rater agreement of the recommendation of EVT were assessed using Gwet’s AC1 (κ_G_) reliability coefficient. AC1 stands for agreement coefficient, first order chance correction and is used for binary data. ^27^ We derived the κ_G_ for binary (EVT recommendations) data, with 95% bias-corrected confidence intervals. The agreement categories were defined as poor (κ_G_ <0), slight (κ_G_ =0–0.20), fair (κ_G_ =0.21–0.40), moderate (κ_G_ =0.41– 0.60), substantial (κ_G_ =0.61–0.80) and excellent (κ_G_ >0.80) according to the Landis and Koch criteria.^28^ Non-overlapping confidence intervals and level of agreement were considered for potential difference between KG values.^29^ The following prespecified subgroup analyses of interrater reliability were conducted: rater specialty, rater experience, infarct core size and time since stoke onset. For intrarater reliability, we looked at rater specialties. We used the R statistical computing language (R Core Team, 2022) in the RStudio framework to compute for the K_G_ values using the irrCAC package. ^30^ The κ_G_ statistic has been shown to be relatively resistant to the Kappa paradox. κ_G_ has a less severe correction for chance agreement compared to Cohen’s Kappa because it does not assume all observed ratings may yield an agreement by chance and it places greater emphasis on hard-to-score subjects. ^27,31^

We had initially planned to perform interrater agreement analysis on the first round of the survey only. However, two new randomized controlled trials (ANGEL-ASPECT and SELECT 2) on EVT for large vessel occlusion stroke patients with large core infarcts were published during the study period.^9,10^ We felt that these new trials might influence how raters make their decisions; thus, we also estimated the interrater reliability on the second round.

### Ethics

The study was approved by the research ethics board of 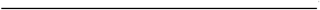.

Consent from patients was waived. Survey participants’ consent was obtained before the survey, and reaffirmed after, to ensure their continued agreement despite the study’s inherent deceptive methodology.

## Results

### Patients

A total of 3,144 cranial CT scans done for symptoms of stroke were reviewed. Of these, 912 were found to have intracranial vascular occlusion. After applying the inclusion criteria, 120 cranial CT scan of large vessel occlusion stroke patients were found eligible (eFigure 1). The final 30 were selected to balance out clinical characteristics and radiologic features.

eTable 1 summarizes the characteristics of the 30 large vessel occlusion stroke patients whose CT scans were selected for final inclusion in the study. The median age was 77.0 years (min, max: 39.0, 92.0), 46.7% were females, most of the occlusions (60%) were on the middle cerebral artery and 40% of the patients underwent treatment with EVT. The mean NIHSS was 17 (standard deviation: 7), the median officially reported ASPECTS was 5 (min, max: 0, 10) and the median core infarct size was 82cc (min, max: 0, 270).

### Raters

Invitations were sent to the members of the two Canadian societies of physicians directly involved in EVT decision-making for large vessel occlusion stroke patients, the Canadian Interventional Neuro Group (31 members) and the Canadian Stroke Consortium (186 members). Thirty-four respondents began responding to the initial survey that opened on November 29, 2022 but 5 did not complete it, leaving 29 respondents available for analysis. Of the 29 that finished the first round of surveys, 11 (37.9%) were stroke neurologists, 11 (37.9%) were neurointerventionalists and 7 were dual trained. Ten (34.5%) were senior raters with 10 or more years of experience. All except one rater finished the second round which opened on January 19, 2023. (See eTable 2)

### EVT Decisions

Involving 60 CT scans of large vessel occlusion stroke cases and 29 raters, a total of 1,740 decisions were rendered regarding the recommendation for or against thrombectomy. The characteristics associated with the decisions regarding treatment made by the raters are summarized in Table 1. Substantial discrepancies in median values and frequencies of recommendations favoring or against EVT are observed in the following: Alberta Stroke Program Early CT Score (ASPECTS) (3 versus 7), core size (125 cc versus 50cc), collateral scores, and National Institutes of Health Stroke Scale (NIHSS) scores (20 versus 14). The percentage of decisions made against (50.7%) or for (49.5%) EVT based on perfusion CT scan was comparable.

**Table 1.** Endovascular thrombectomy decisions.

| Characteristic | Decision: No EVT<br>(N=724) | Decision: EVT<br>(N=1016) | Overall<br>(N=1740) |
| --- | --- | --- | --- |
| NIHSS: Median [Min, Max] | 20.0 [6.00, 27.0] | 14.0 [6.00, 27.0] | 17.5 [6.00, 27.0] |
| Laterality of occlusion: Left | 222 (30.7%) | 474 (46.7%) | 696 (40.0%) |
| Time from stroke onset to presentation<br>(Hours): Median [Min, Max] | 13.1 [6.95, 21.5] | 13.1 [6.95, 21.5] | 13.1 [6.95, 21.5] |
| Sex: Female | 337 (46.5%) | 475 (46.8%) | 812 (46.7%) |
| Age (Years): Median [Min, Max] | 79.0 [39.0, 92.0] | 75.0 [39.0, 92.0] | 77.0 [39.0, 92.0] |
| Specialty of Rater |  |  |  |
| Stroke Neurology | 315 (43.5%) | 375 (36.9%) | 690 (39.7%) |
| Neurointervention | 261 (36.0%) | 399 (39.3%) | 660 (37.9%) |
| Dual trained | 148 (20.4%) | 242 (23.8%) | 390 (22.4%) |
| Experience of Rater (Years): Median<br>[Min, Max] | 4.00 [2.00, 33.0] | 4.00 [2.00, 33.0] | 4.00 [2.00, 33.0] |
| Imaging presented at each decision |  |  |  |
| Perfusion CT neuroimaging | 367 (50.7%) | 503 (49.5%) | 870 (50.0%) |
| Non-perfusion CT neuroimaging | 357 (49.3%) | 513 (50.5%) | 870 (50.0%) |
| ASPECTS given by the rater:<br>Median [Min, Max] | 3.00 [0, 10.0] | 7.00 [1.00, 10.0] | 5.00 [0, 10.0] |
| Infarct core size<br>Median [Min, Max] | 125 [0, 270] | 50.0 [0, 270] | 81.0 [0, 270] |
| Collateral grading given by the rater |  |  |  |
| 0: Absence of vessels on CTA distal<br>to occlusion | 146 (20.2%) | 38 (3.7%) | 184 (10.6%) |
| 1: Collateral supply filling 0-50% of<br>MCA territory | 405 (55.9%) | 266 (26.2%) | 671 (38.6%) |
| 2: Collateral supply filling 51-99% of<br>MCA territory | 150 (20.7%) | 437 (43.0%) | 587 (33.7%) |
| 3: Collateral supply filling 100% of<br>MCA territory | 23 (3.2%) | 275 (27.1%) | 298 (17.1%) |

NIHSS National institutes of health stroke score; SD Standard deviation; CT Computed tomography ASPECTS Alberta stroke program early CT score

The mosaic plot in Figure 1 shows the distribution of EVT decisions for 870 cases (30 cases shown to 29 raters twice—non-perfusion CT neuroimaging and perfusion CT neuroimaging). The overall recommendation for EVT was 59.0% (513/870) without perfusion images and 57.8% (503/870) with perfusion images a difference of only 1.1%. The observed agreement for no EVT (30.9%, 269/870) and for EVT (47.7%, 415/870) was 78.6% (684/870). Among 870 cases studied, 11.3% (98 cases) recommended EVT based on non-perfusion CT but not on perfusion CT scans, while 10.1% (88 cases) advised against EVT on non-perfusion CT but recommended it on perfusion CT for the same patients. Despite the small difference 1.1% in total EVT recommendations, decisions changed in 21.4% of cases.

**Figure 1.**
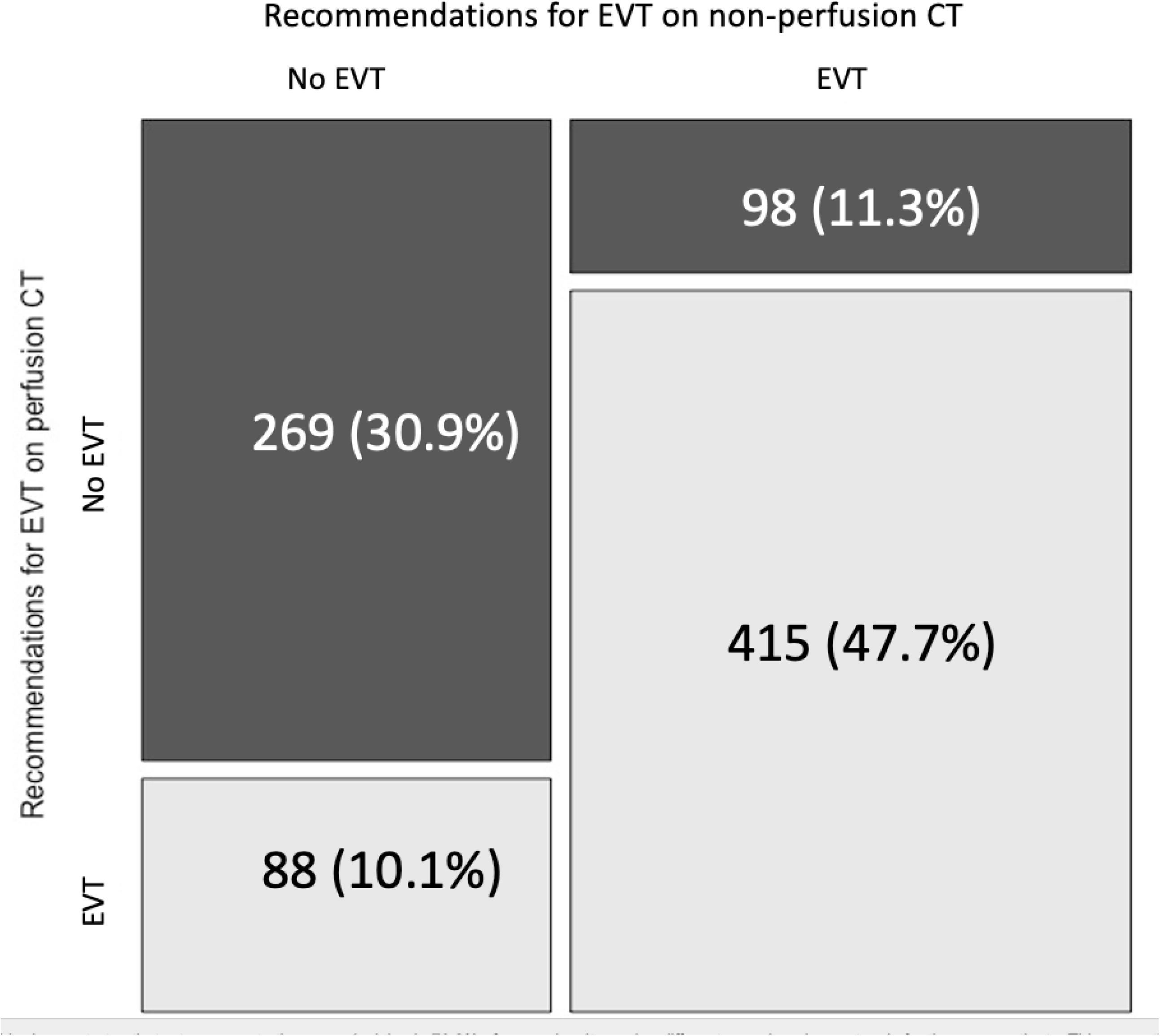
How does neuroimaging affect endovascular thrombectomy (EVT) decisions? A mosaic plot with a contingency table overlay is utilized to display the distribution of 870 cases (30 patients x 29 raters) and the corresponding decisions for or against endovascular thrombectomy (EVT) between two imaging protocols. All percentages are based on the denominator of 870. All box sizes are proportional to the magnitude of decisions. For instance, the lower left box represents the 10.1% (88/270) of all decisions. In this box, EVT was not recommended on non-perfusion imaging but recommended on perfusion imaging. Conversely the upper right quadrant of the plot signifies cases where EVT was recommended based on non-perfusion neuroimaging but not on perfusion neuroimaging (11.3%, 98/270). Conversely, the lower left quadrant represents cases that were not recommended for EVT based on non-perfusion CT neuroimaging but were deemed appropriate for EVT following perfusion CT neuroimaging.

### Inter-rater Agreement

The interrater (κ_G_) agreement is similar with non-perfusion CT neuroimaging compared to perfusion CT neuroimaging (κ_G_ = 0.487; 95% CI 0.327,0.647 and κ_G_ =0.552; 95% CI 0.430,0.675). (See Figure 2) The confidence intervals overlap and both K_G_ signify moderate agreement.^28^ Similarly the interrater agreement was also not significantly different between the neuroimaging protocols when calculated according to physician subspecialty and experience. Both the subgroups for dual trained physicians and small core infarcts achieved substantial agreement across both neuroimaging protocols. The level of agreement increased (moderate to substantial) from non-perfusion CT neuroimaging to perfusion CT neuroimaging in the subgroups of stroke onset of 6-12 hours CT (κ_G_ =0.502; 95% CI 0.237,0.767and κ_G_ =0.621; 95% CI 0.427,0.815) and large core infarcts (κ_G_ = 0.451; 95% CI 0.367,0.76 and κ_G_ =0.665; 95% CI 0.411,0.919). (See Table 2)

**Figure 2.**
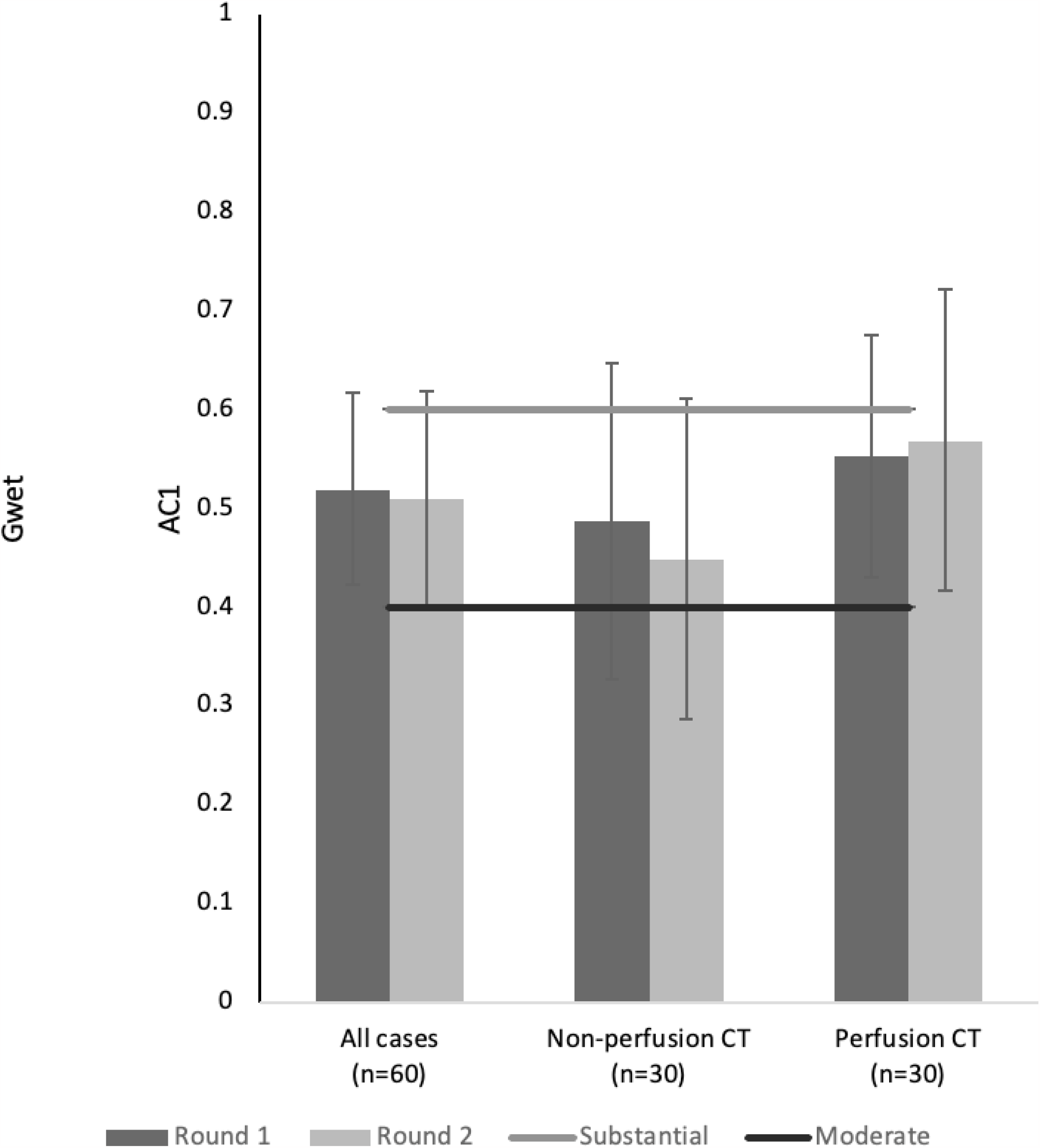
Interrater Agreement for Endovascular Thrombectomy Between Two Surveys. Clustered bar graphs with error bars show Gwet’s AC1 (κ_G_) statistic with 95% confidence interval for inter-rater agreement between neuroimaging modalities and two survey rounds. There is extensive confidence intervals overlap between neuroimaging modalities and time from stroke onset indicating no differences. The level of agreement is marked as moderate (lower black line) or substantial (higher gray line) above the threshold lines.

**Table 2.**
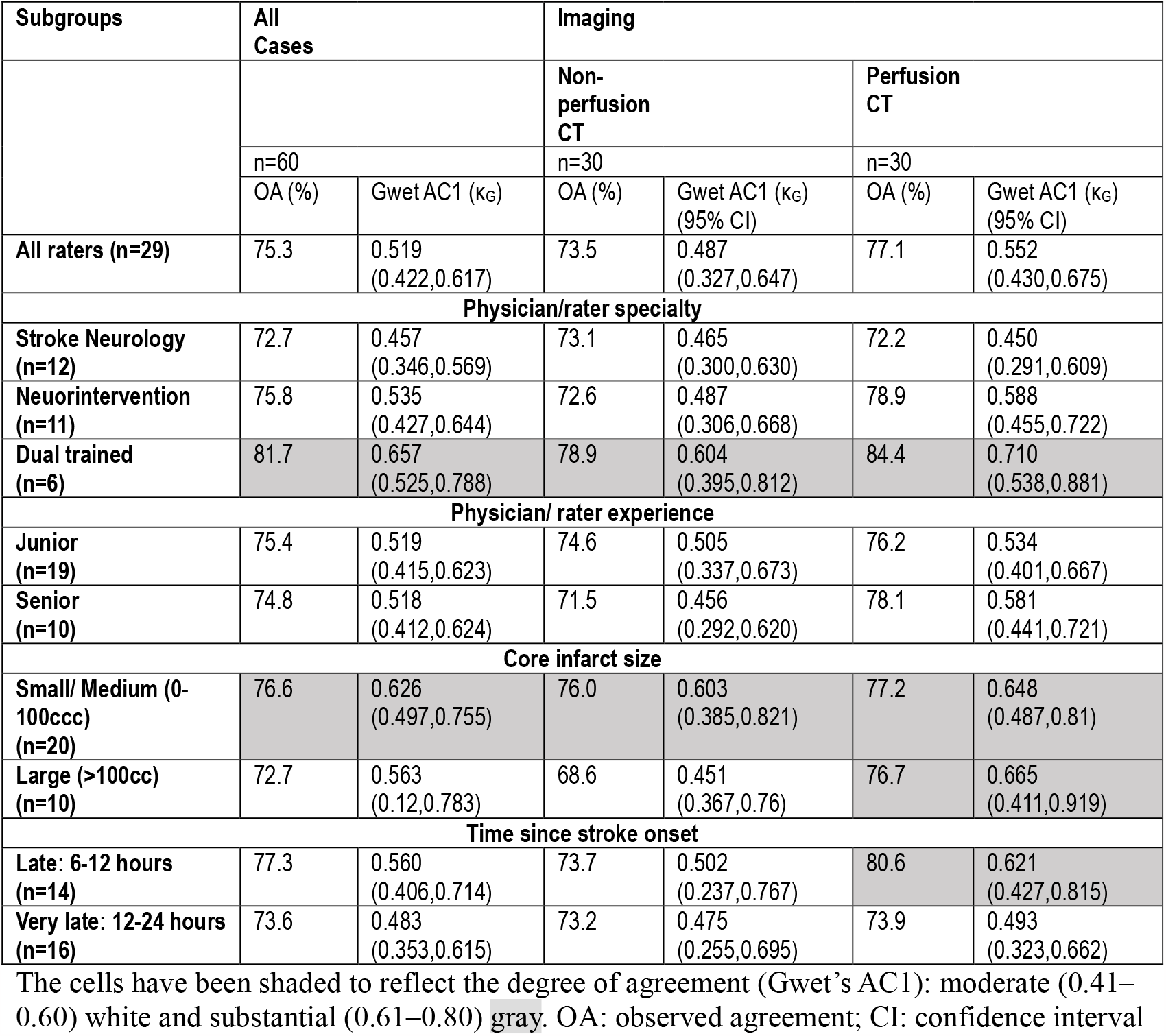
Interrater Agreement for Endovascular Thrombectomy.

As we elaborated on earlier, we performed an unplanned inter-rater analysis of the second survey round. (See Figure 2) There was no significant increase or decrease in agreement between the first and second round in both neuroimaging protocols as confidence intervals overlapped and stayed in the moderate agreement classification. Even for the subgroup of large core infarcts, the agreement level stayed at moderate in the second round (κ_G_ = 0.443; 95% CI 0.081,0.805 and κ_G_ = 0.550; 95% CI 0.253,0.847).

### Intrarater Reliability

Twenty-eight (out of 29) raters finished both surveys. Table 3 summarizes the intra-rater agreement of these 28 raters according to the type of neuroimaging reviewed for their decisions. Confidence intervals of all intra-rater agreements overlap between both neuroimaging protocols for each of the 28 raters signifying no significant difference as per our predefined criteria.

**Table 3.** Intra-rater agreement for Endovascular Thrombectomy.

|  | Overall |  | Non-perfusion CT |  | Perfusion CT |  |
| --- | --- | --- | --- | --- | --- | --- |
| Rater ID | OA (%) | Gwet's AC1 (95% CI) | OA (%) | AC1 (95% CI) | OA (%) | AC1 (95% CI) |
| 1 | 80.0 | 0.602<br>(0.393,0.810) | 73.3 | 0.469<br>(0.133,0.805) | 86.7 | 0.735<br>(0.477,0.992) |
| 2 | 86.7 | 0.741<br>(0.566,0.915) | 86.7 | 0.738<br>(0.482,0.994) | 86.7 | 0.744<br>(0.49,0.997) |
| 3 | 83.3 | 0.694<br>(0.506,0.882) | 86.7 | 0.760<br>(0.516,1.00) | 80.0 | 0.627<br>(0.327,0.926) |
| 4 | 83.3 | 0.679<br>(0.488,0.871) | 83.3 | 0.694<br>(0.42,0.968) | 83.3 | 0.670<br>(0.388,0.952) |
| 5 | 90.0 | 0.800<br>(0.644,0.956) | 90.0 | 0.800<br>(0.572,1) | 90.0 | 0.802<br>(0.575,1) |
| 6 | 81.7 | 0.673<br>(0.480,0.867) | 86.7 | 0.792<br>(0.572,1) | 76.7 | 0.546<br>(0.225,0.867) |
| 7 | 86.7 | 0.804<br>(0.66,0.948) | 86.7 | 0.804<br>(0.594,1) | 86.7 | 0.804<br>(0.594,1) |
| 8 | 78.3 | 0.599<br>(0.387,0.811) | 83.3 | 0.706<br>(0.438,0.974) | 73.3 | 0.487<br>(0.149,0.825) |
| 9 | 91.7 | 0.858<br>(0.73,0.986) | 90.0 | 0.816<br>(0.601,1) | 93.3 | 0.896<br>(0.742,1) |
| 10 | 76.7 | 0.580<br>(0.363,0.797) | 73.3 | 0.562<br>(0.239,0.885) | 80.0 | 0.615<br>(0.314,0.917) |
| 11 | 80.0 | 0.671<br>(0.479,0.864) | 73.3 | 0.540<br>(0.21,0.87) | 86.7 | 0.792<br>(0.572,1) |
| 12 | 86.7 | 0.738<br>(0.562,0.914) | 83.3 | 0.676<br>(0.395,0.956) | 90.0 | 0.801<br>(0.575,1) |
| 13 | 76.7 | 0.627<br>(0.422,0.831) | 80.0 | 0.689<br>(0.418,0.959) | 73.3 | 0.562<br>(0.239,0.885) |
| 14 | 86.7 | 0.744<br>(0.57,0.917) | 80.0 | 0.627<br>(0.327,0.926) | 93.3 | 0.869<br>(0.682,1) |
| 15 | 80.0 | 0.602<br>(0.393,0.81) | 90.0 | 0.800<br>(0.572,1) | 70.0 | 0.416<br>(0.065,0.768) |
| 16 | 81.7 | 0.680<br>(0.489,0.871) | 83.3 | 0.733<br>(0.482,0.985) | 80.0 | 0.627<br>(0.327,0.926) |
| 17 | 81.7 | 0.650<br>(0.451,0.849) | 70.0 | 0.416<br>(0.065,0.768) | 93.3 | 0.876<br>(0.696,1) |
| 18 | 80.0 | 0.604<br>(0.396,0.812) | 83.3 | 0.670<br>(0.388,0.952) | 76.7 | 0.538<br>(0.217,0.859) |
| 19 | 85.0 | 0.701<br>(0.515,0.887) | 83.3 | 0.670<br>(0.388,0.952) | 86.7 | 0.733<br>(0.475,0.992) |
| 20 | 80.0 | 0.655<br>(0.457,0.853) | 73.3 | 0.540<br>(0.210,0.870) | 86.7 | 0.770<br>(0.533,1) |
| 21 | 92.0 | 0.849<br>(0.714,0.983) | 90.0 | 0.817<br>(0.601,1) | 93.3 | 0.880<br>(0.705,1) |
| 22 | 70.0 | 0.416<br>(0.175,0.657) | 66.7 | 0.359<br>(-0.006,0.724) | 73.3 | 0.491<br>(0.139,0.813) |
| 23 | 78.3 | 0.568<br>(0.353,0.782) | 80.0 | .602<br>(0.298,0.905) | 76.7 | 0.546<br>(0.225,0.867) |
| 24 | 80.0 | 0.621<br>(0.414,0.827) | 70.0 | 0.450<br>(0.097,0.802) | 90.0 | 0.805<br>(0.581,1) |
| 25 | 71.6 | 0.485<br>(0.249,0.721) | 66.7 | 0.378<br>(0.01,0.745) | 76.7 | 0.589<br>(0.275,0.902) |
| 26 | 86.7 | 0.736<br>(0.56,0.912) | 86.7 | 0.751<br>(0.502,1) | 86.7 | 0.735<br>(0.477,0.992) |
| 27 | 88.3 | 0.768<br>(0.602,0.935) | 83.3 | 0.667<br>(0.384,0.95) | 93.3 | 0.872<br>(0.688,1) |
| 28 | 88.3 | 0.768<br>(0.599,0.934) | 83.3 | 0.667<br>(0.384,0.95) | 93.3 | 0.867<br>(0.678,1) |

The cells have been shaded to reflect the degree of agreement (Gwet's AC1): Fair (0.21–0.40) green moderate (0.41–0.60) white, substantial (0.61–0.80) gray and excellent (>0.80) black OA: observed agreement

Seventeen of the 28 raters (60.7%) had a change in the classification of intrarater agreement from non-perfusion CT neuroimaging to perfusion CT neuroimaging. With the addition of automated CT perfusion images, 11/17 had improved by at least one category while 6/17 had worsened by at least one category. eFigure 2 demonstrates the distribution of K_G_ for intra-rater reliability of the 28 raters. It illustrates that while the K_G_ values remain in the same category (substantial), the median value for perfusion CT is higher than non-perfusion CT.

## Discussion

Our study pursues two interconnected yet distinct objectives. Concerning decisions related to EVT. we observed a change in approximately 21.4% of decisions when perfusion imaging was incorporated. In terms of reliability, our findings revealed no statistically significant disparities in both intra- and interrater reliability, whether perfusion imaging was included or not. The propensity of raters to arrive at consistent decisions for cases remained comparable across both groups.

### EVT Decisions

Even though the raters were looking at the same patient clinical data, NCCTs and CTAs, their decisions for EVT changed in one out of every 5 patients (21.4%, 186/870) with perfusion imaging and these changes occurred in both directions, 10.1% (88/870) for EVT and 11.3% (98/870) against EVT. Taken together these changes in decisions with perfusion CT neuroimaging only led to 1.1% (10/870) fewer recommendations for EVT. Tabulating EVT recommendations, we saw that decisions for EVT were more common for patients with better collaterals, higher ASPECTS, smaller infarcted cores. These are all intuitive as these factors all portend a better prognosis to large vessel occlusion stroke patients undergoing EVT.^25,32^

There are several possible reasons for these changes in decisions based on the neuroimaging presented. For those that go from recommending EVT to recommending against it (non-perfusion CT neuroimaging to perfusion CT neuroimaging), this may be from seeing the core infarct quantified. While early ischemic changes may not be apparent on non-contrast computed tomography (NCCT), they may appear larger on automated perfusion images. This could be attributed to genuine ischemia or the manifestation of the ghost core phenomenon, wherein the core infarct appears larger on perfusion imaging than its actual size. ^33^ (See Patient 7 in eFigure 3) For those that go from recommending against EVT to recommending it (non-perfusion CT neuroimaging to perfusion CT neuroimaging), the decision is likely based on seeing a large value for the penumbra on automated perfusion images or a smaller core to the ASPECTS they calculated. Despite the possibility that the frank hypodensity observed on non-contrast computed tomography (NCCT) is the only region affected by the arterial occlusion, automated perfusion images can reveal viable regions surrounding the core infarct that could potentially be salvaged. (See Patient 17 in eFigure 3)

### Interrater Reliability

Interrater agreement (K_G_) on the decision to recommend EVT for late window large vessel occlusion stroke patients was comparable whether raters viewed non-perfusion CT neuroimaging or perfusion CT neuroimaging. Both were classified as moderate agreement and overlapping confidence intervals. The same results, overlapping confidence intervals at the same level of agreement, were found in both the first and second rounds of the survey. These findings support our hypothesis that the addition of perfusion imaging does not alter the reliability of decisions made by physicians of different specialties on a wide spectrum of large vessel occlusion stroke patients. In contrast, a similar study showed overlapping confidence intervals with the addition of automated perfusion imaging but had an increase in agreement from moderate (K = 0.506) to substantial (K=0.704) with the addition of automated perfusion imaging.^34^ This is likely accounted for by the differences in study design as the aforementioned study utilized only two raters and gave an “indecisive” option for EVT decisions.

### Intrarater Reliability

The intrarater agreement statistics (κ_G_) on the decision to recommend EVT for late window large vessel occlusion stroke patients for each rater had overlapping confidence intervals between non-perfusion CT neuroimaging or perfusion CT neuroimaging. The extremes of intra-rater agreement are striking: non-perfusion CT (min, max: 0.359, 0.817) and perfusion CT (min, max: 0.416, 896). We posit that this difference may be due to some reviewers already being familiar with the newly published randomized trials and some not.^10,11^ In essence, the reviewers with lower intrarater agreement may have “disagreed” with themselves on account of the new evidence. While those with higher K_G_ may not have reviewed the evidence or do not yet subscribe to it.

### Limitations

Our study has notable limitations, including the absence of a universally accepted method for comparing agreement statistics (KG) beyond magnitude documentation. To gauge similarity in KG values, we relied on overlapping confidence intervals and comparable levels of agreement. ^29^ Additionally, the study’s applicability is restricted to Canadian-based raters, and it does not encompass other stroke imaging modalities. We also omitted detailed clinical information and did not conduct an analysis of clinical outcomes in relation to the chosen imaging modality. Finally, examining causality between baseline patient and rater variables, including the choice of neuroimaging modality, and the decision to recommend EVT is not within the study’s scope.

## Conclusion

In this study, we found that the addition of CTP presented to the raters altered 21.4% of the decisions on EVT but the net difference in EVT recommendations was only 1.1% (10/870) between the two neuroimaging protocols. Secondly, the study suggests that addition of perfusion imaging to NCCT and CTA does not significantly affect the interrater and intrarater reliability of the decisions made by physicians on EVT for late window large vessel occlusion stroke patients. Stated differently, the propensity of raters arriving at consistent decisions for the same cases, whether in comparison to other raters (interrater) or their own assessments (intrarater), remains similar regardless of the presence or absence of perfusion imaging in the review process.

Stated differently, the propensity of raters arriving at consistent decisions for the same cases, whether in comparison to other raters (interrater) or their own assessments (intrarater), remains similar regardless of the presence or absence of perfusion imaging in the review process.

## Data Availability

The full data is available upon reasonable request from the authors.

## Abbreviations

CTA: Computed tomography angiography
NCCT: Non-contrast cranial CT
EVT: Endovascular thrombectomy
LVO: large vessel occlusion
ASPECTS: Alberta Stroke Program Early CT Score

## Notes

### Competing Interest Statement

JBD: Honoraria from Medtronic. Travel grant from Microvention; MAM: Member of the scientific advisory board of Palmera Medical; TRM: Principal of eVasc Neurovascular;

### Funding Statement

JBD is a recipient of the Fergus Mills Scholarship of McMaster University for the Clinical Uncertainty in Large Vessel Occlusion (CULVO) study.

### Author Declarations

The study was approved by the research ethics board of Unity Health- St. Michael's Hospital (SMH REB# 22-186).

